# Oxytocin and its role in caloric intake and appetite: A preregistered living systematic review and meta-analysis

**DOI:** 10.64898/2026.03.25.26349278

**Authors:** Alina I. Sartorius, Elisabeth Deilhaug, Heemin Kang, Damien Dufour, Kjersti Mæhlum Walle, Kamryn T. Eddy, Dennis van der Meer, Lars T. Westlye, Ole A. Andreassen, Elizabeth A. Lawson, Daniel S. Quintana

## Abstract

Oxytocin is a hypothalamic hormone and neuromodulator that has been linked to a variety of different functions, including parturition, social behavior, and cognitive processing. More recently, oxytocin has also been associated with metabolism and energy balance. However, evidence to date in this field has been inconsistent, especially in human research. To address this, we performed a preregistered systematic review and meta-analysis, which synthesized existing literature on the effect of exogenous oxytocin administration compared to a placebo on caloric intake and appetite in humans, using a living meta-analysis approach. Results indicated a significant, reductive effect of oxytocin administration on appetite in participants belonging to certain patient groups (e.g., obesity or type II diabetes; Hedges’ *g* = −0.21). A separate moderator analysis evaluating oxytocin’s effect on caloric intake revealed a conditional effect depending on the patient group, with the obesity group showing a significant effect. We did not find any statistically significant effects in healthy participants. However, further analyses revealed that these effects were also not equivalent, indicating that the data are currently too insensitive to draw clear conclusions. Taken together, the results provide some evidence for the role of oxytocin in regulating appetite in an anorexigenic, possibly homeostatic fashion. Future updates in this living meta-analysis may lead to more definitive conclusions.

## Introduction

The neuromodulator and hormone oxytocin is a versatile nonapeptide that is mainly synthesized in the supraoptic and paraventricular (PVN) nuclei of the hypothalamus [1], alongside vasotocin (also known as (arginine) vasopressin or antidiuretic hormone) [2, 3]. The hypothalamus plays a critical role in energy balance and metabolism [4, 5]. The hypothalamic PVN specifically contribute to the regulation of feeding via excitatory and inhibitory inputs to neuron pathways in the hypothalamic arcuate nuclei, which are essential for hunger-driven feeding [5], and the hypothalamus contains ‘glucose sensing’ neurons which are thought to be involved in mediating glucose homeostasis [6, 7].

Oxytocin is well-known for its role in uterine tissue contractions during childbirth and its essential role in the milk let-down reflex [8–10]. In the late 1990s and early 2000s, research emerged demonstrating its contribution to cognitive functions and social behavior, which remains a topic of considerable research interest to date [11, 12]. Evidence has also been accumulating indicating that oxytocin may be involved in aspects of energy balance, energy regulation, and metabolism [13–17], next to its complex social and cognitive properties. Energy homeostasis can be conceptualized as a continuous internal monitoring of an organism comparing energy intake (e.g., via ingestion) versus energy expenditure (e.g., via movement or thermoregulation). Energy balance denotes the difference between intake and expenditure [18]. If an otherwise healthy organism has a negative energy balance, that is, it lacks energy (and might in the long-term be eventually underweight), hunger signaling and food-seeking behavior should be increased. If it has a positive energy balance (surplus of energy, possible overweight), hunger signaling and food-seeking behavior should be decreased. Of note, hunger and appetite cues are not the only indicators and controls of food intake and energy balance, since feeding behavior and energy homeostasis are highly complex biological mechanisms (see [19]). In obesity for instance, homeostatic mechanisms can be altered (e.g., leptin resistance [20]). Metabolism is tightly connected to energy balance and encompasses a suite of biochemical reactions and processes through which an organism sustains itself, and regulates energy storage and use [21, 22].

There are several ways through which the effects of oxytocin on energy regulation and metabolism can present themselves and be measured, such as through changes in body weight and composition, appetite and food intake, regulation of biomarkers or neuronal pathways, or measurement of energy expenditure. Most early studies investigating the neurobiological mechanistic pathways of oxytocin were conducted in animal models. These revealed that lesions to the PVN, including oxytocinergic projections, can cause excessive food consumption (i.e., hyperphagia) ([23, 24]; in rodents), implying that hypothalamic oxytocin circuits are likely involved in the regulation of feeding behavior. Intracerebroventricular (ICV) oxytocin injections decreased food intake [15, 25] and body weight, possibly mediated by upregulated lipolysis in adipose tissue, indicating that fat metabolism could be implicated, at least in rodents [26, 27]. Deblon and colleagues also found that oxytocin could increase blood glycerol and decrease triglyceride levels, and have a protective effect on insulin resistance [27]. Endogenous hypothalamic oxytocin has been suggested to play a role in leptin-regulated food intake mechanisms in rats [28]. Oxytocin’s role in appetite regulation is further supported by the distribution of *OXTR* (the gene encoding the oxytocin receptor) mRNA in brain regions also related to appetite and feeding [29–31].

In humans, research has predominately explored how oxytocin regulates appetite and caloric intake. Compared to measures of body composition and biomarkers, the presence or absence of appetite and food intake behavior are accessible measures of a neutral, negative, or positive balance of energy [18]. These variables are relatively easy to assess via self-report and do not require longitudinal follow-up — as would be necessary for the monitoring of body weight changes for instance — or laboratory testing. Importantly, they are also non-invasive, in contrast to for instance severing of the PVN in non-human animals. The physiological feeling of appetite, hunger, satiety, and fullness are among many key feedback signals to control food intake, which in turn is indicative of increased or decreased energy requirements [4]. Notably, appetite and food cravings can also be hedonic and not necessarily signify energy imbalance [32]. Nevertheless, resulting changes in ingestion behavior can impact energy balance.

For instance, Striepens and colleagues (2016) reported that a single administration of intranasal oxytocin led to a reduction in food cravings in healthy, typical-weight adult women compared to a placebo [33]. Another study by Thienel and colleagues (2016) found that single-dose intranasal oxytocin provoked a significant decrease in food intake in obese, but otherwise healthy men, in comparison to a placebo condition [34]. Alongside this finding, further studies link intranasal oxytocin to a reduction in homeostatic and reward-driven food intake in humans [35–38]. Conversely, Leslie and colleagues found no significant differences between exogenous oxytocin and placebo on food intake in healthy individuals and individuals with bulimia or binge eating disorder (BED), however, oxytocin reduced hunger in patients with bulimia and/or BED [39].

Studies have also investigated the relationship between metabolic biomarkers and body weight and oxytocin in humans. Oxytocin’s immediate role in energy metabolism is inherently complex to study, not least because ‘metabolism’ in itself is a broad term that may refer to a myriad of underlying pathways. On a biological level, this can translate into among other lipolysis (briefly mentioned above; the process of breaking down triglycerides which releases fatty acids and glycerol [40]), or into proxy biomarkers of metabolism such as plasma (or blood) glucose, triglyceride, insulin, leptin, and ghrelin levels, and many other measures [18]. In human males, a single dose of intranasal oxytocin (compared to a placebo) has been linked to insulin suppression and reduced glucose concentrations after meal-intake in typical-weight but not men with obesity [41]. In terms of the link between oxytocin and weight loss or body composition, both positive [42, 43] and non-significant effects [38, 44] have been reported. Espinoza and colleagues could show that intranasal oxytocin was linked to an increase in lean body mass in individuals with sarcopenic obesity [42]. At the same time, McCormack and colleagues did not find an effect of intranasal oxytocin on body weight in individuals with hypothalamic obesity [44].

Taken together, current evidence indicates that oxytocin could play a role in energy balance and metabolism, however, the evidence to date has been inconclusive. To our knowledge, no formal meta-analysis on this topic has been conducted in the past five years. In 2021, one team of researchers conducted a systematic review and meta-analysis investigating the effects of intranasal oxytocin on food intake and cravings in non-psychiatric individuals, overweight individuals, and individuals with an eating disorder [45]. However, in their review, the authors did not include psychiatric patients, did not specify hunger or satiety outcomes in their search string, and did not account for effect size dependency, which can bias results since multiple effects were extracted from some studies [46]. In addition, the review did not assess publication bias, which can inflate effect sizes, leading to incorrect conclusions regarding the summary effect size. Publication bias refers refers to the tendency to omit the publication of certain study outcomes depending on the strength or direction of the effect [47]. Indeed, there is evidence for publication bias in the disciplines of psychology and medicine, including oxytocin research [48, 49]. Lastly, the review did not include physiological indicators of food or energy needs such as feelings of hunger or satiety. Leslie and colleagues reported a meta-analysis on oxytocin and energy intake in 2018. However, new results have amounted in the meantime, such that an updated, more comprehensive analysis with updated sensitivity tools is warranted [50]. Other reviews that have been performed included data synthesis components on endogenous oxytocin and eating behavior, metabolism, appetite regulation, and food intake, however, no formal meta-analysis was conducted and no data synthesis on exogenously administered oxytocin was performed (e.g., [29, 51]).

To address this gap in synthesized data in the literature, we performed a preregistered living systematic review and meta-analysis evaluating the link between the effects of exogenous oxytocin on different markers of caloric intake and appetite, using recently developed methods to account for publication bias. While meta-analyses remain the gold standard for synthesizing evidence on a topic [52], they can quickly become outdated due to the rapid publication of new studies [53, 54], especially in the field of oxytocin [11]. To counteract this issue, this meta-analysis is ‘living’ [55, 56], in that it is planned to be updated twice within a timeline of the upcoming four years, as specified in our study protocol [57]. Living meta-analyses expand and improve the original concept of a meta-analysis. With new studies being added to an original meta-analysis project on a regular basis, the validity of the claims of the meta-analysis is maintained, and conclusions become more robust since they are based on a steadily growing and renewing pool of effects. This increases power and sensitivity of the data. We conducted four subgroup meta-analyses to capture the full complexity of aspects that constitute appetite and caloric intake, and to assess oxytocin’s effects in both healthy and clinical individuals. The meta-analyses were grouped into subgroups based on a pre-liminary scoping review of the literature: a) caloric intake in healthy individuals, b) caloric intake in clinical individuals, c) appetite in healthy individuals, and d) appetite in clinical individuals. To better understand how different covariates, such as sex, age, study design, or administration dose and frequency, may influence the variation in results, we included moderator analyses. The original protocol for this meta-analysis is available on PROSPERO (https://www.crd.york.ac.uk/PROSPERO/view/CRD420251025864; see supplementary document 1 for deviations).

## Methods

### Literature search and screening

The literature search was performed following the Preferred Reporting Items for Systematic Reviews and Meta-Analyses (PRISMA) 2020 statement for Living Systematic Reviews (PRISMA-LSR) [58]. The searches were performed on April 7th, 8th, and 10th, 2025. Search terms were determined in collaboration with a local academic librarian (University of Oslo Library), defined in the analysis protocol and used as such ([57]; also see supplementary document 7), and customized to each database. The following databases were searched: PubMed/Medline, Embase, APA PsycInfo, Cochrane Library, Scopus, and Web of Science. All screening of the studies was handled in Covidence (https://www.covidence.org/). The first author (AIS) independently screened the titles and abstracts of all studies for initial eligibility, and the full-texts of all potentially eligible studies for further selection. Co-author ED cross-checked the full-texts of a random 25% subsample of these studies. Disagreements were resolved by senior author DSQ. Studies were considered eligible if they met the following inclusion criteria: a) Individuals from both healthy (as defined in the primary study) and clinical populations (e.g., obesity, anorexia nervosa, schizophrenia (SCZ)), b) exogenous administration of oxytocin (including syntocinon, excluding carbetocin) via the intranasal, intravenous, oral, or intramuscular route, c) within- or between-participant randomized placebo-controlled trials to study the effect of oxytocin, d) outcome measures related to energy balance and metabolism operationalized through biological markers, caloric intake and appetite, and body weight/composition, and e) the study is available in English. Studies were considered ineligible and excluded if they were a) animal studies, b) investigating the effects of endogenous oxytocin, or c) cohort studies, case reports, case-control studies, or cross-sectional studies. For caloric intake, we considered studies investigating any type of caloric, food, energy, or volume intake or consumption, feeding, and hyperphagia. For appetite, we considered related markers such as hunger, satiety, fullness, (hedonic) drive to eat, and food craving (see protocol [57] for the full search string). The majority of duplicates and ineligible/non-RCT studies were automatically marked by Covidence. Remaining duplicates were manually identified by AIS in Covidence. At this point of the process, the meta-analysis deviated from the protocol due to resource constraints. The planned secondary search to identify more potentially eligible studies was not performed. Furthermore, two (*biomarkers* and *body composition*) of the three originally planned sub-meta-analyses were not performed and we proceeded with the ‘markers of *caloric intake* and *appetite*’ meta-analysis only. This meta-analysis in turn was handled as four separate sub-analyses, one each focusing on outcomes related to caloric intake in healthy and clinical cohorts, and one each focusing on outcomes related to appetite in healthy and clinical cohorts.

### Data extraction

To calculate the standardized mean differences and their variances (i.e., Hedges’ *g*) between oxytocin and placebo conditions for the different outcome measures of caloric intake and appetite, data was extracted from each study by first-author AIS. Summary data (i.e., means, standard deviations (SD), standard errors (of the mean; SE or SEM), group sample sizes (N)) could be extracted for all or some relevant outcome measures for 17 studies [34–39, 42, 59–68]. WebPlotDigitizer [69] was used to retrieve summary data estimates from two studies [37, 59]. The variances (i.e., standard deviation or standard error) from these extracted means were estimated by subtracting the outermost error bar value from the mean point value. The reliability and precision of WebPlot-Digitizer was validated using an eligible study [63] with both summary data and plots available (see example in previously published meta-analysis [70, 71]; supplementary table 2). For 19 studies [17, 34–36, 39, 44, 59, 66, 67, 72–81], summary data for all or some outcome measures was not readily extractable, thus, anonymized raw or summary data was requested directly from the authors. Raw or summary data was received from six studies [17, 34, 36, 72, 74, 78], leading to the complete exclusion of eight studies from analyses [44, 73, 75–77, 79–81]. In addition to the summary data, study characteristics such as publication year, study design, oxytocin administration frequency, population demographics, and study outcomes were collected (supplementary table 1).

In three studies [17, 72, 78], oxytocin and placebo were infused continuously via an intravenous line, and outcomes (i.e., satiety, volume intake) were measured continuously alongside. In these cases, three options were followed: a) the outcomes at the time point closest to 15-20 minutes after start of infusion were chosen since intravenous oxytocin infusions have been shown to be detectable in cerebrospinal fluid after approximately 15 minutes ([82], in macaques), and central satiety signals peak around 15 to 25 minutes after meal ingestion [83] (for study [78]), b) the maximum satiety score relative to the baseline score was calculated (for studies [17, 72]), and/or c) the satiety score 30 minutes after terminating a meal relative to the baseline score was calculated (for studies [17, 72]). In any case, it was tried to mimic to the methodological approach used in the primary study as closely as possible. In two studies [34, 74], some or all of the outcome measures were recorded and intended to be interpreted over five hours (measured across six time points), thus, the mean and SD for the area under the curve (AUC) were calculated as outcomes for the oxytocin and placebo conditions. Standard errors were converted to standard deviations. For chronic administration studies where a baseline and end-of-treatment values were reported (e.g., [62]), the end-of-treatment values for the oxytocin and placebo conditions were preferably extracted for the sake of consistency. Alternatively, if only the differences between baseline and end-of-treatment was reported, this value was utilized. Due to resource constraints, co-author HK did not extract data from a random 25% sample of the eligible studies to assess inter-rater reliability of data extraction, as originally specified in the protocol. Instead, co-author DD validated all data extracted by AIS, but did not extract data separately.

### Statistical analyses

If not stated otherwise, all statistical analyses were conducted with the statistical software R (version 4.5.2; [84]) in RStudio (version 2025.09.2+418; [85]). The R package `tidyverse` (version 2.0.0; [86]) was used for core data handling (see below and supplementary document 8 for further R packages used). The following analysis steps were performed in the healthy and clinical cohorts separately and also separately for the *caloric intake* and the *appetite* meta-analyses, with the exception of the effect size calculations, which were performed in bulk for i) all effects for caloric intake outcomes and ii) all effects for appetite outcomes.

#### Effect size calculations

For the calculation of effect sizes, we used the R package `metafor` (version 4.8.0; [87]). Effect sizes for within-participant/repeated measures design studies were generated using the standardized mean change with change score standardization (SMCC; [88]). For a precise calculation of effect sizes and their variances, we calculated Pearson’s *r* as the correlation coefficient which measures the correlation between the oxytocin and placebo responses within participants, based on the raw data and summary data we received [17, 34, 36, 72, 74, 78] and found in online supplementary information [66]. The average correlation coefficient (*r*_Caloric intake_ = 0.7679153 and *r*_Appetite_ = 0.5774734) was used for the remaining outcomes where no raw data or coefficient was available. For between-participant studies, the standardized mean difference (SMD) in the form of Hedges’ *g* was calculated. Hedges’ *g* was chosen as the main effect size measure since sample sizes between studies varied considerably [89, 90], and Hedges’ *g* can correct for small sample bias. For both SMCC and SMD calculations, means, SD and N were used for calculations, and standard errors (of the mean) were converted into SD.

After the initial calculations, effect sizes were coded as follows. An increase in caloric intake and appetite/hunger/drive to eat, as well as less satiety and fullness after oxytocin administration versus placebo were coded as ‘positive’, and conversely, a decrease in caloric intake and appetite/hunger/drive to eat, as well as more satiety and fullness were coded as ‘negative’. This means that, for instance, in a study where a cohort consumed more calories in the oxytocin condition compared to the placebo condition and the calculated effect size had a negative sign, the sign was reversed to positive (the calculated effect sizes and their variances are reported in supplementary table 3). In summary, in all participants, the coding represents an increase in energy intake as positive and a decrease in energy intake as negative.

#### Meta-analyses

Since many of the included studies reported multiple outcome measures and thus, multiple effect sizes were extracted from the same study, a frequentist three-level random-effects meta-analysis was performed to account for variability between effect sizes within studies [91]. This meta-analysis was performed using the R package `metafor` (version 4.8.0; [87]). A Bonferroni-adjusted *α*-threshold of 0.017 (0.05 divided by three) was used for the evaluation of statistical significance of the main meta-analysis to account for multiple comparisons due to the two planned prospective analyses, as this is a living meta-analysis with repeated testing [57]. The frequentist multi-level approach was complemented with a robust hierarchical Bayesian model-averaged meta-analysis (hBMA). The Bayesian approach comes with the possibility of quantification of evidence for the null and alternative hypothesis, whereas a traditional frequentist approach can only allow for rejection or failure to reject the null hypothesis [92]. For this, the R package `RoBMA` (version 3.6.0; [93]) was used. The resulting Bayes factor was interpreted as follows: Bayes factor <1 is considered evidence favoring the null hypothesis, Bayes factor 1-3 is interpreted as anecdotal evidence favoring the alternative hypothesis, Bayes factor >3 is interpreted as moderate evidence, and Bayes factor >10 as strong evidence for the alternative hypothesis [94].

#### Equivalence testing and heterogeneity assessment

It is conceivable that the calculated effect sizes do not reach statistical significance although a true effect may be underlying and present. This can be due to for instance underpowered analyses [95]. Thus, frequentist equivalence tests were performed to explore this possibility and to gauge whether the statistical non-significance of effect sizes is due to the absence of a meaningful effect or due to insensitive data, using the R package `TOSTER` (version 0.8.6; [96]). Equivalence testing allows for the rejection of effects that are larger than a ‘smallest effect size of interest’ (SESOI) [97]. The equivalence bounds were informed with the smallest effect sizes as determined by [98] (based on different meta-analyses of oxytocin intervention studies corrected for publication bias). Therefore, two SESOIs were selected: more stringent bounds of Hedges’ *g* = ± 0.06 and more relaxed bounds of Hedges’ *g* = ± 0.23, given what effect sizes can be observed in the oxytocin literature [70, 95, 98].

Within-study heterogeneity can be caused by variances in latent participant variables such as weight or multiple effects from the same study measuring marginally different aspects of the same concept. Between-study heterogeneity stems from variability across studies for instance in experimental designs. Heterogeneity between studies and within studies can influence the size of the calculated effects and the summary effect size. To quantify the presence and degree of heterogeneity regarding outcome measures, the ‘Restricted Maximum Likelihood’ (REML) [99] option integrated in the frequentist meta-analysis model was used, which yields the *Q* measure. Additionally, we assessed between- and within-study heterogeneity with the *I*^2^-statistic, which is based on the two multilevel random-effects meta-analysis variance components 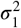 (between-cluster heterogeneity) and 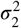 (within-cluster heterogeneity) [100].

#### Power analysis and bias assessments

For the statistical power analysis, the package `metameta` (version 0.2; [101]) was used. `metameta` allows for the calculation of study-level power for meta-analyses for a range of hypothetically possible effect sizes of interest for all included studies, given that the true effect size of a population is usually unknown. The potential presence of publication bias was estimated with a sensitivity analysis, using the R package `PublicationBias` (version 2.4.0; [102]). The use of robust Bayesian meta-analysis to complement the other publication bias measure was not feasible since the publication bias correction in robust hierarchical Bayesian model-averaged meta-analysis has not been validated yet [103]. Small study bias (indicative of publication bias) was assessed using Egger’s regression test [102] implemented in the R package `meta` (version 8.2-1; [104]) and visualized with a contour-enhanced funnel plot [105]. In contrast to the pre-registration protocol, the tool `RoB 2` [106] to assess general risk of bias in randomized trials was not deployed, due to resource constraints.

#### Moderator analyses

Moderator analyses were conducted to estimate the degree of unexplained between-study heterogeneity that may be accounted for by covariates or moderator variables, respectively. Covariates were defined *a priori* in the preregistered study protocol as (1) study design (within-versus between-participants), (2) study population (clinical versus healthy), (3) sex (percentage female participants), (4) age, (5) administration route, (6) dosage (<24 International Units (IU), 24 IU, >24 IU), (7) administration frequency (chronic (i.e., > 2 doses) versus non-chronic (i.e., ≤ 2 doses)), (8) endpoint measure (objective versus subjective reporting), (9) endpoint status (primary versus secondary outcome), and (10) peer review status. In the course of the data extraction, however, due to the structure of the data and amendments to the analysis plan, the moderator analyses had to be adjusted. Since healthy and clinical populations were analyzed separately, moderator (2) was dropped from all analyses. The following moderators were removed since they contained only one level and no variation in the respective meta-analysis sample: Moderator (1) and (7) for MA1, moderator (10) for MA2, moderator (1), (7) and (8) for MA3, and moderator (10) for MA4. Furthermore, we added two moderators post-hoc. A moderator indicating whether the outcome had been measured fasted or non-fasted (moderator (11)). The condition type was added as a moderator to replace the original moderator (2), assessing the impact of the condition (i.e., whether someone had obesity, SCZ, etc.) on the summary effect size (moderator (12)).

## Results

### Literature search and screening

The search yielded an initial sample of 12986 records, of which 4803 studies were screened. K = 2766 records were removed as duplicates (96.6% marked by Covidence) and k = 5417 records were automatically removed by Covidence due to ineligibility/non-RCT study design. After title and abstract screening of the remaining 4803 studies, 4535 records were removed because they did not meet the inclusion criteria a) to e) and/or did meet the exclusion criteria a) to c). 268 studies remained and underwent full-text screening. AIS and ED disagreed on the status of five studies, which was resolved by DSQ and resulted in 52 studies being initially identified as eligible for data extraction. During the data extraction process, another four studies were identified as not eligible and excluded (k = 48). Due to only conducting the meta-analyses on *markers of caloric intake and appetite*, the number of eligible studies was further reduced to k = 33. Following this, four additional studies were excluded during the data extraction stage due to irrelevance of outcome, leading to a sample of k = 29 studies. Of the 239 studies excluded, 66 could not be retrieved (i.e., original record to available or accessible). Reasons for exclusion for the remaining 173 studies were irrelevant outcome (k = 68), non-RCT design (k = 25), no placebo condition (k = 20), full-text not in English (k = 18), studies removed due to resource constraints (k = 15), use of animal model (k = 14), measurement of endogenous oxytocin (N = 9), and no administration of exogenous oxytocin (k = 4). Finally, eight studies were excluded, despite eligibility for inclusion, due to data unavailability. Thus, the final sample of studies being included in the analyses was k = 21 (***Figure 1***; supplementary document 2 and supplementary table 1 for the included studies).

**Figure 1.**
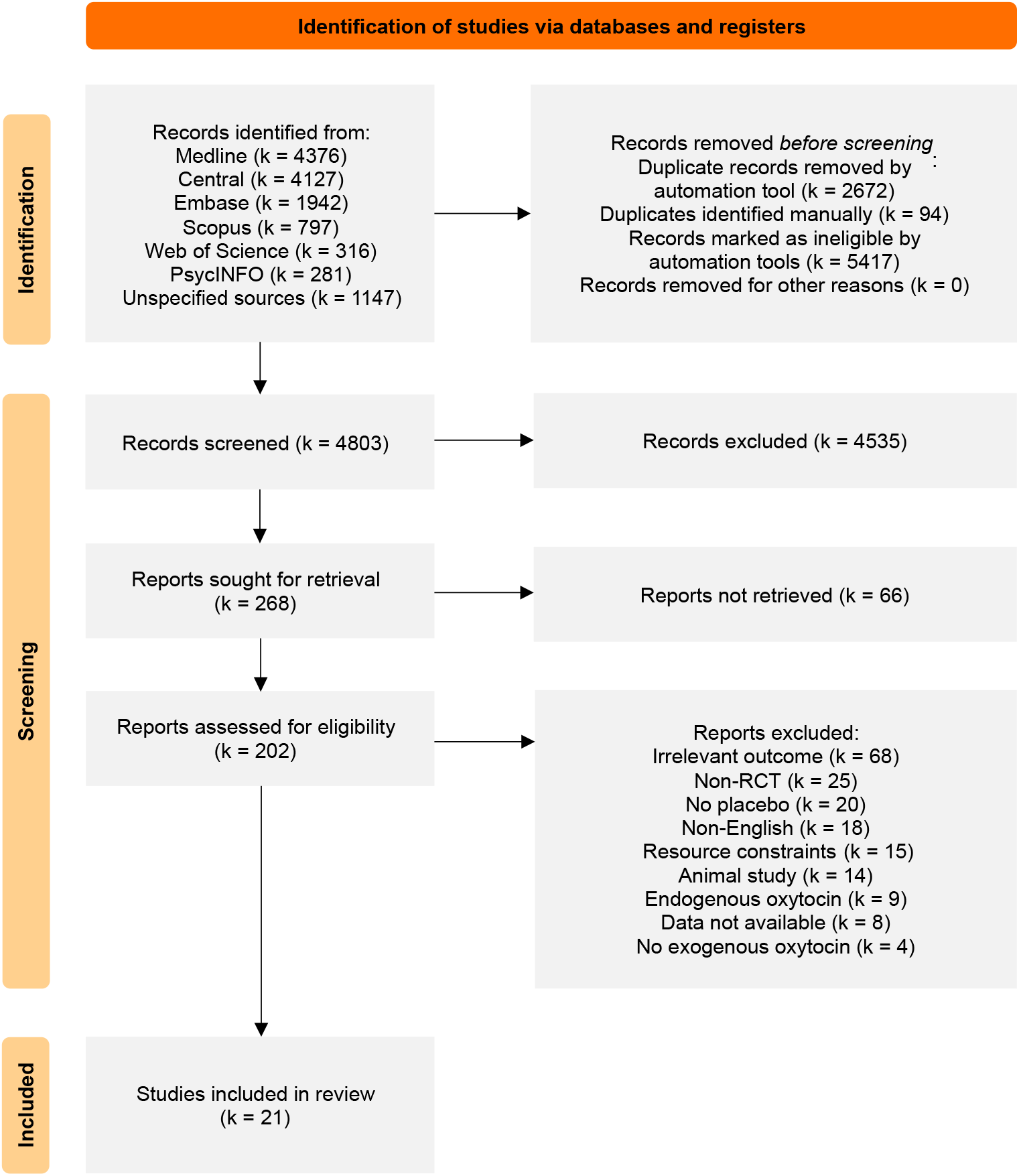
PRISMA flow diagram. ‘Resource constraints’ refers to the studies that were eligible for the originally additional two planned meta-analyses on biomarkers and measures of body composition, but were removed due to resource constraints (see *Methods* section). RCT, randomized clinical trial.

### Study characteristics

Across the 21 studies, 691 individuals were included. The mean age across studies was 31.16 years (SD = 12.96 years). 18 studies had a within-participant design (n = 597 participants) and three a between-participant design (n = 94 participants). n = 356 individuals had a clinical condition (anorexia nervosa: 96, obesity: 90, Prader-Willi syndrome: 44, bulimia only: 34, bulimia or BED: 25, T2D: 25, SCZ: 16, functional dyspepsia: 14, diabetic gastroparesis: 12) and n = 335 were healthy. The majority of studies (k = 18 [34–39, 42, 59–68, 74]) used the intranasal route for the administration of the oxytocin/placebo, otherwise, oxytocin/placebo were intravenously infused [17, 72, 78]. Two thirds of the included ‘intranasal’ studies administered 24 IU [34–38, 42, 59, 60, 66, 68, 74, 107], five administered more than 24 IU [39, 61, 63–65] and one study used less than 24 IU [62]. Four studies evaluated the effects of three or more repeated administrations (i.e., chronic administration [17, 38, 42, 61, 62]) while 17 used a non-chronic scheme with two or less administrations [34–37, 39, 59, 60, 63–68, 72, 74, 78]. For a detailed overview of study characteristics, see supplementary table 1.

### Meta-analyses

Four frequentist multilevel meta-analyses with three levels were performed, using the restricted maximum likelihood estimator (REML) [87] to account for potential effect size dependencies due to multiple effects being extracted from some studies. An adjusted *α*-threshold of 0.017 was applied to correct for future living meta-analysis updates [57]. The meta-analysis on measures of caloric intake in healthy individuals (MA1) included 19 effect sizes across ten studies and yielded a summary effect size of Hedges’ *g* = −0.225 [CI −0.436, −0.015] (*p* = 0.036). The meta-analysis on the same outcome in clinical individuals (MA2) was performed with 16 effects across nine studies and produced a summary effect size of Hedges’ *g* = −0.049 [CI −0.234, 0.137] (*p* = 0.607; ***Figure 2***). The two meta-analyses on measures of appetite were performed on 18 effect sizes across nine studies in healthy individuals and on ten effects across seven studies in clinical individuals. In the healthy population (MA3), this produced a summary effect size of Hedges’ *g* = 0.074 [CI −0.137, 0.284] (*p* = 0.492). In the clinical population (MA4), the resulting summary effect size was Hedges’ *g* = −0.208 [CI −0.362, −0.054] (*p* = 0.008; ***Figure 3***), indicating that oxytocin reduces appetite in clinical populations. See supplementary documents 3 − 6 for detailed meta-analysis results.

**Figure 2.**
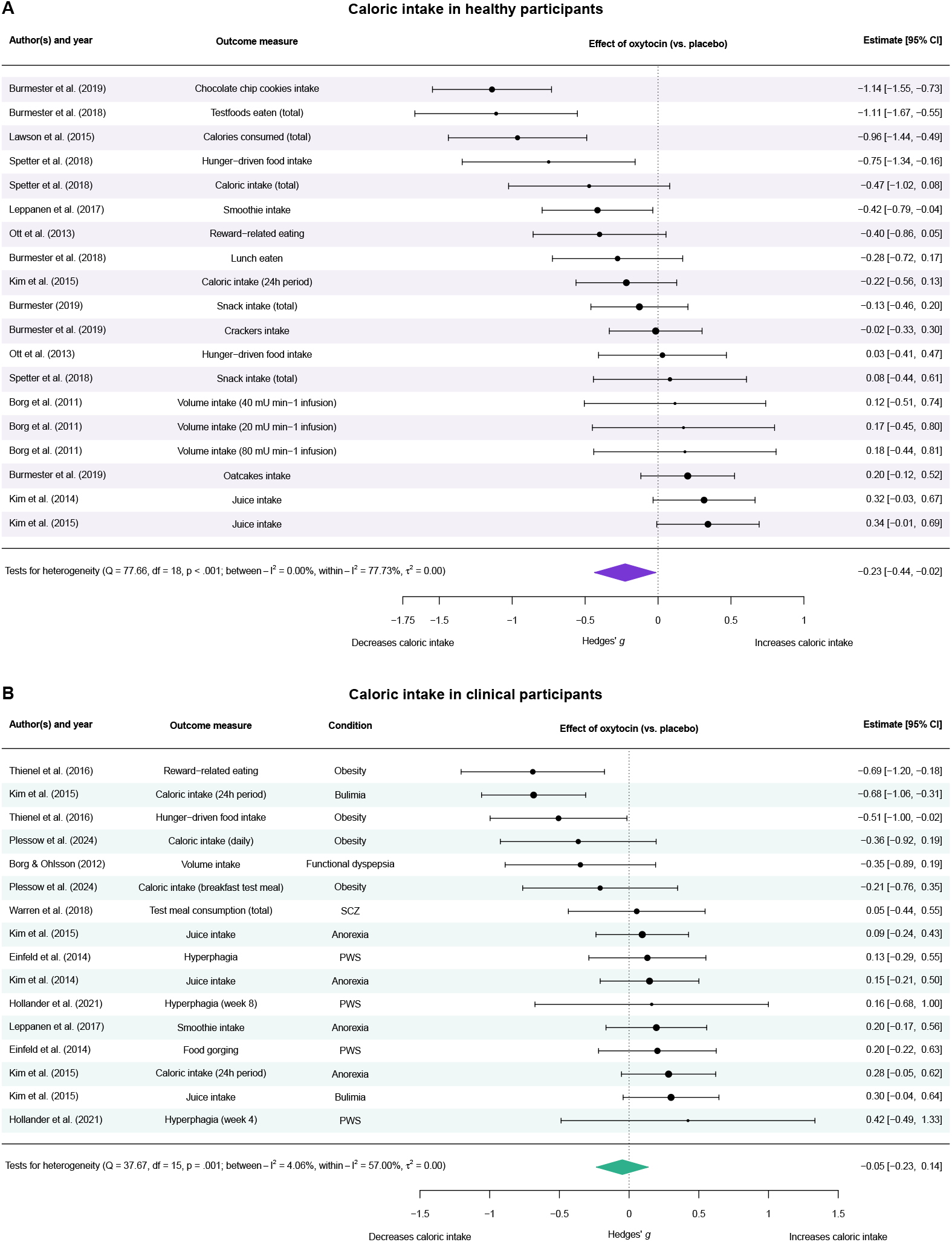
Forest plots for caloric intake in healthy and clinical participants. (A) Distribution of effect sizes for 19 outcomes across ten studies in ascending order. The study’s first author and publication year are displayed in the outer left column, followed by the measured outcome, effect sizes (black dots) and their variances (whiskers), and the exact estimates including their 95% confidence interval. The summary effect size (Hedges’ *g* = −0.225) is displayed at the bottom of the plot as a purple diamond. min-1, min^-1^. (B) Distribution of effect sizes for 16 outcomes across nine studies in ascending order. The study’s first author and publication year are displayed in the outer left column, followed by the measured outcome, the clinical condition, effect sizes (black dots) and their variances (whiskers), and the exact estimates including their 95% confidence interval. The summary effect size (Hedges’ *g* = −0.049) is displayed at the bottom of the plot as a teal diamond. *General notes*. Point sizes of the effect sizes are relative and a function of the model weights. The vertical dotted line indicates a Hedges’ *g* = 0. CI, confidence interval; PWS, Prader-Willi syndrome; SCZ, schizophrenia. Compare to data in supplementary table 3 and supplementary documents 3 - 4.

**Figure 3.**
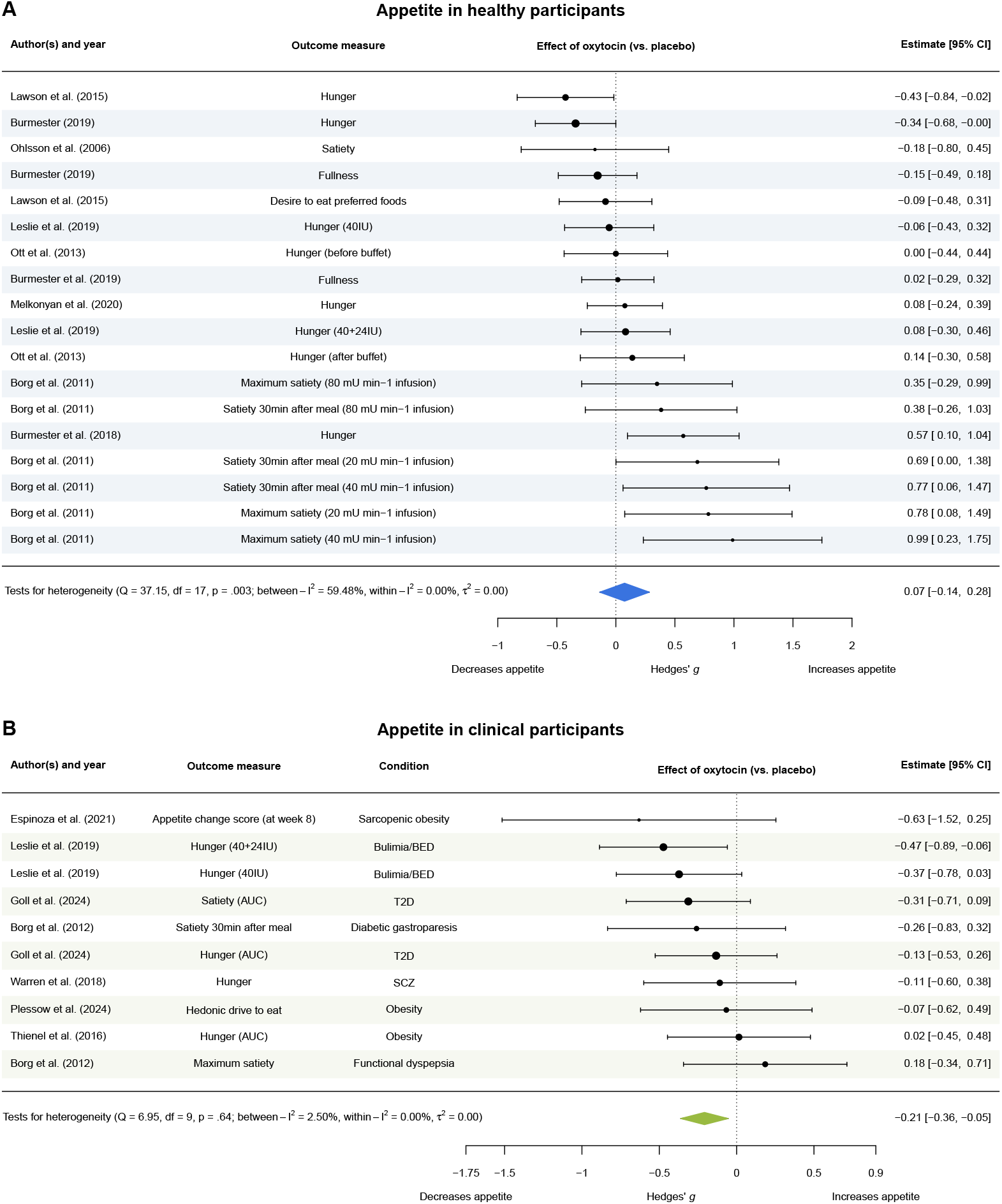
Forest plots for appetite in healthy and clinical participants. (A) Distribution of effect sizes for 18 outcomes across nine studies in ascending order. The study’s first author and publication year are displayed in the outer left column, followed by the measured outcome, effect sizes (black dots) and their variances (whiskers), and the exact estimates including their 95% confidence interval. The summary effect size (Hedges’ *g* = 0.074) is displayed at the bottom of the plot as a blue diamond. min-1, min^-1^. (B) Distribution of effect sizes for ten outcomes across seven studies in ascending order. The study’s first author and publication year are displayed in the outer left column, followed by the measured outcome, the clinical condition, effect sizes (black dots) and their variances (whiskers), and the exact estimates including their 95% confidence interval. The summary effect size (Hedges’ *g* = −0.208) is displayed at the bottom of the plot as an olive green diamond. *General notes*. Point sizes of the effect sizes are relative and a function of the model weights. The vertical dotted line indicates a Hedges’ *g* = 0. CI, confidence interval; T2D, type II diabetes mellitus. Compare to data in supplementary table 3 and supplementary documents 5 - 6.

Each frequentist meta-analysis was complemented with a Bayesian hierarchical meta-analysis. For MA1 (caloric intake in healthy participants), the posterior probability was 0.409 and the inclusion BF 0.692 (*μ*_*mean*_ = −0.089 [CI −0.395, 0.000]). For MA2 (caloric intake in clinical participants), the posterior probability was 0.095 with an inclusion BF of 0.105 (*μ*_*mean*_ = −0.004 [CI −0.107, 0.020]). In MA3 (appetite in healthy participants), the posterior probability was 0.113 and the inclusion BF was 0.127 (*μ*_*mean*_ = 0.008 [CI 0.000, 0.147]). Therefore, the BFs in MA1 - MA3 correspond to evidence favoring the null hypothesis. Lastly, for MA4 (appetite), the posterior probability was 0.646 and the inclusion BF was equal to 1.823 (*μ*_*mean*_ = −0.133 [CI −0.350, 0.000]), indicating anecdotal evidence for the alternative hypothesis.

### Equivalence testing and measures of heterogeneity of studies and samples

We performed equivalence tests to check whether the results we reported above, especially the statistically non-significant results (i.e., MA1, MA2, MA3), are due to statistical equivalence (i.e., a truly trivial effect). Effect sizes were converted to Cohen’s *d* for compatibility with the R package `TOSTER`. For MA1, using the conservative equivalence bounds of *d* = ± 0.06, the equivalence test was non-significant, meaning the null equivalence hypothesis cannot be rejected [*Z* = −1.568 (*p* = 0.942)]. For the equivalence bounds of *d* = ± 0.23, the equivalence test was also not significant [*Z* = −0.0437 (*p* = 0.517)]. For MA2, the null equivalence hypotheses with equivalence bounds of ± 0.06 [*Z* = 0.096 (*p* = 0.462)] could not be rejected, however, the null equivalence hypotheses with equivalence bounds of *d* = ± 0.23 could be rejected, i.e., the equivalence test was significant [*Z* = 1.827 (*p* = 0.034)]. Finally, for MA3, the equivalence tests with the equivalence bounds of *d* = ± 0.06 and ± 0.23 both were non-significant [*Z* = 0.176 (*p* = 0.570) and *Z* = −1.320 (*p* = 0.093)].

Regarding study and sample heterogeneity, the frequentist multilevel meta-analyses indicated the presence of heterogeneity for the majority of meta-analyses (MA1: *Q*(df = 18) = 77.661, *p* < .001; MA2: *Q*(df = 15) = 37.674, *p* = 0.001; MA3: *Q*(df = 17) = 37.148, *p* = 0.003), except for MA4 (Q(df = 9) = 6.948, *p* = 0.643). The more detailed *I*^2^-statistics revealed that, for MA1 (caloric intake in healthy participants), 1.36e-7% of the total variance stems from between-study heterogeneity and 77.73% from within-study heterogeneity. For MA2 (caloric intake in clinical participants), 4.06% of the total variance can be attributed to between-study heterogeneity and 56.99% to within-study heterogeneity. In MA3 (appetite in healthy participants), the *I*^2^-statistics calculation showed that 59.48% of the variance can be attributed to between-study and only 6.46e-9% to within-study heterogeneity. For MA4, 2.50% of the variance could be attributed to between-study and 2.44e-6% to within-study heterogeneity.

### Power of included studies

The calculations of statistical power for the four separate meta-analyses revealed different median power values to detect the four observed summary effect sizes. For the caloric intake meta-analysis in healthy individuals (MA1, Hedges’ *g* = −0.225), the observed power was 16.74%. For the same outcome in clinical individuals (MA2), the observed power to detect the effect size (Hedges’ *g* = −0.049) was 5.51% (***Figure 4***). For the meta-analysis on markers of appetite in the healthy individuals (MA3), the power for the effect size of Hedges’ *g* = 0.074 was at 6.25%, and for clinical individuals(MA4) at 13.73% with a Hedges’ *g* of −0.208 (***Figure 4***).

**Figure 4.**
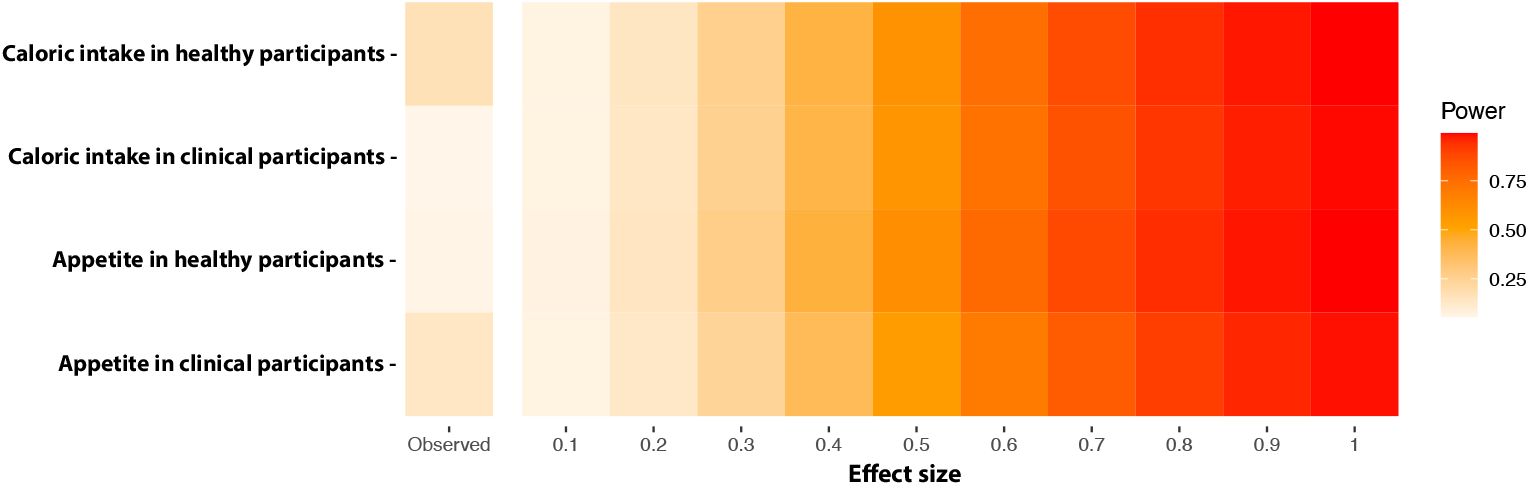
Fireplots for MA1 - MA4. Displayed are the fireplots for each meta-analysis indicating the observed summary effect size of the respective meta-analysis (left outer most column) and its power for a range of ‘true’ effect sizes and their power (aggregated columns to the right). MA1 is displayed at the top row, followed by MA2, MA3 and MA4 in the bottom row. The raw data underlying this plot is presented in supplementary tables 4 - 5.

### Bias assessments

After correcting for publication bias using a robust selection model, the summary effect sizes shrank for all meta-analyses, however, none of the changes were statistically significant. For MA1, Hedges’ *g* shrank from −0.225 to −0.162 [CI −0.369, 0.046 (*p* = 0.109)]. Hedges’ *g* decreased from −0.049 to −0.006 [CI −0.212, 0.201 (*p* = 0.946)] in MA2. Similarly, it marginally decreased in size for MA3 from 0.074 to *g* = 0.068 [CI −0.255, 0.390 (*p* = 0.630)]. For MA4, the sum of effects decreased after adjusting for publication bias from −0.208 to Hedges’ *g* = −0.197 [CI − 0.411, 0.0166 (*p* = 0.061)].

We also tested for small study bias and publication bias for all four meta-analyses using Egger’s regression test and contour-enhanced funnel plots for additional visual inspection. For the two meta-analyses on measures of caloric intake, the funnel plots (***Figure 5***A-B) appear near-symmetric, with a mild skew at the bottom of ***Figure 5***B. This visual impression is reflected in Egger’s regression test results, which did not yield significant results (MA1: t(17) = −1.36, *p* = 0.192, bias estimate −2.779 (SE = 2.046); MA2: t(14) = −1.02, *p* = 0.326, bias estimate −1.580 (SE = 1.553)). The funnel plot for MA3 (***Figure 5***C) shows more visible asymmetry, with studies with large standard errors and effects accumulating to the right bottom of the plot, while the left bottom is relatively emptier. This is confirmed with the result from the regression test, which provides significant evidence for funnel plot asymmetry (t(16) = 3.94, *p* = 0.001, bias estimate 3.573 (SE = 0.907)). The funnel plot for MA4 (***Figure 5***D) shows no clear sign of asymmetry, corresponding to the non-significant Egger’s test result (t(8) = 0.13, *p* = 0.897, bias estimate 0.197 (SE = 1.471).

**Figure 5.**
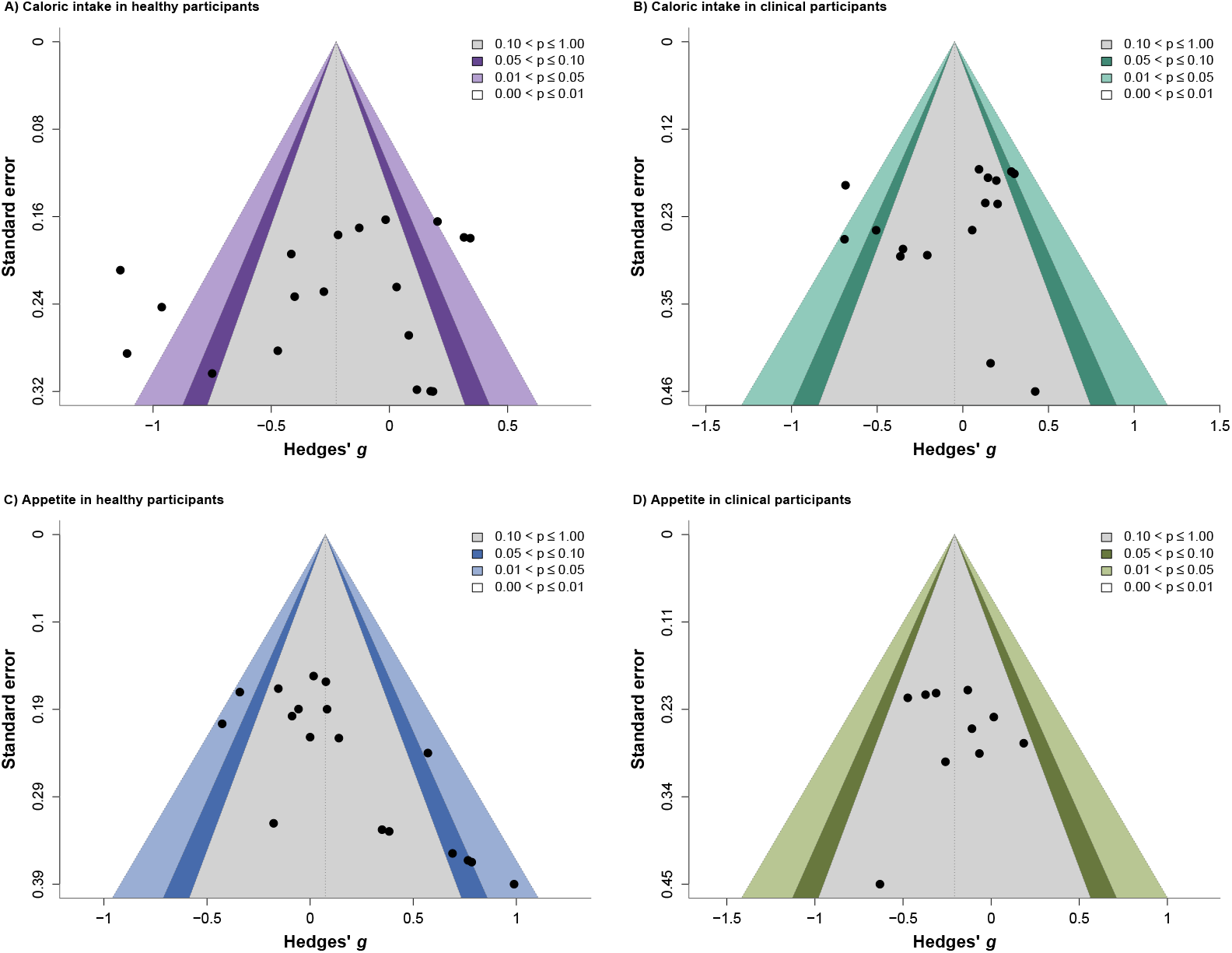
Contour-enhanced funnel plots for the four meta-analyses. Each black dot represents an effect size for one outcome from a study. The vertical dotted lines represent the summary effect size; the contour-shaded areas signify *p*-value cutoffs (see legends). (A) Plot for the meta-analysis on caloric intake in healthy individuals. (B) Plot for the meta-analysis on caloric intake in clinical individuals. (C) Plot for the meta-analysis on appetite in healthy individuals. (D) Plot for the meta-analysis on appetite in clinical individuals.

### Moderator analyses

For MA1, MA3 and MA4, none of the moderators had a statistically significant effect. In MA2 (caloric intake in clinical participants), three moderators were significant. The variable ‘sex’ significantly moderated the pooled effect size (QM(df = 1) = 4.209, *p* = 0.040). The estimated effect was more positive in samples with a higher percentage of female participants (*β* = 0.590 [CI 0.026, 1.154], SE = 0.288, *p* = 0.040) and was more negative in samples with no female participants (*β* = −0.541 [CI −1.027, −0.054], SE = 0.248, *p* = 0.029). For the same meta-analysis, the oxytocin dosage had a significant effect on the outcome (QM(df = 2) = 6.020, *p* = 0.049). The pooled effect was significantly more negative at 24 IU (*β* = −0.342 [CI −0.649, −0.036], SE = 0.156, *p* = 0.029) and shifted towards a more positive effect at doses >24 IU relative to 24 IU (*β* = 0.429 [CI 0.060, 0.797], SE = 0.188, *p* = 0.023). Doses <24 IU did not differ significantly from 24 IU. A post-hoc moderator analysis assessing the effect of the condition type (i.e., which diagnosis a clinical group had) had a significant effect on the pooled outcome of MA2 (QM(df = 5) = 11.842, *p* = 0.037). In relation to the reference group (Prader-Willi syndrome (PWS)), the obesity group showed a significantly more negative effect (*β* = −0.649 [CI −1.129, −0.169], SE = 0.2451, *p* = 0.008). In MA4, the condition type was not significant (QM(df = 6) = 6.389, *p* = 0.381) (see supplementary document 4). An overview of the results from all moderator analyses is presented in Table 1.

**Table 1.** Results from the moderator analyses.

| Meta-analysis (MA#) | Moderator | # | QM (df) | p-value |
| --- | --- | --- | --- | --- |
| Caloric intake in healthy participants (MA1) | % Female | 3 | 2.698 (1) | 0.101 |
|  | Mean age | 4 | 1.077 (1) | 0.299 |
|  | Administration route | 5 | 1.959 (1) | 0.162 |
|  | Oxytocin dose | 6 | 2.389 (1) | 0.122 |
|  | Endpoint measure | 8 | 0.000 (1) | 0.984 |
|  | Endpoint status | 9 | 0.732 (1) | 0.392 |
|  | Peer-review status | 10 | 0.049 (1) | 0.824 |
|  | Meal timing | 11 | 0.057 (2) | 0.972 |
| Caloric intake in clinical participants (MA2) | Study design | 1 | 0.008 (1) | 0.930 |
|  | % Female | 3 | 4.209 (1) | **0.040** |
|  | Mean age | 4 | 0.539 (1) | 0.463 |
|  | Administration route | 5 | 0.611 (1) | 0.435 |
|  | Oxytocin dose | 6 | 6.019 (2) | **0.049** |
|  | Administration frequency | 7 | 0.307 (1) | 0.579 |
|  | Endpoint measure | 8 | 0.074 (1) | 0.786 |
|  | Endpoint status | 9 | 0.058 (1) | 0.810 |
|  | Meal timing | 11 | 0.934 (1) | 0.334 |
|  | Condition type | 12 | 11.842 (5) | **0.037** |
| Appetite in healthy participants (MA3) | % Female | 3 | 0.084 (1) | 0.772 |
|  | Mean age | 4 | 0.726 (1) | 0.394 |
|  | Administration route | 5 | 2.739 (1) | 0.098 |
|  | Oxytocin dose | 6 | 0.005 (1) | 0.946 |
|  | Endpoint status | 9 | 0.000 (1) | 0.996 |
|  | Peer-review status | 10 | 1.403 (1) | 0.237 |
|  | Meal timing | 11 | 1.341 (1) | 0.247 |
| Appetite in clinical participants (MA4) | Study design | 1 | 0.013 (1) | 0.910 |
|  | % Female | 3 | 0.735 (1) | 0.391 |
|  | Mean age | 4 | 1.108 (1) | 0.293 |
|  | Administration route | 5 | 1.094 (1) | 0.296 |
|  | Oxytocin dose | 6 | 2.133 (1) | 0.144 |
|  | Administration frequency | 7 | 0.013 (1) | 0.910 |
|  | Endpoint measure | 8 | 0.254 (1) | 0.615 |
|  | Endpoint status | 9 | 0.022 (1) | 0.882 |
|  | Meal timing | 11 | 0.743 (1) | 0.389 |
|  | Condition type | 12 | 6.389 (6) | 0.381 |

Notes. MA#, Meta-analysis number; #, Moderator number; QM, Q-test for moderators; df, degrees of freedom. Significant moderators are printed in **bold**. Compare to data in supplementary documents 3 - 6.

## Discussion

Here we performed a systematic review and meta-analysis to evaluate the effects of exogenous oxytocin on caloric intake and appetite as behavioral markers of metabolism and energy balance in humans. A total of 63 effect sizes extracted from 21 studies were included across four separate meta-analyses. Out of the four meta-analyses we performed (i.e., markers of caloric intake in (1) healthy and (2) clinical participants, markers of appetite in (3) healthy and (4) clinical participants), MA4 revealed a significant summary effect size of Hedges’ *g* = −0.208, indicating that exogenous oxytocin administration is associated with decreased appetite in the included clinical populations. The other meta-analyses did not yield a statistically significant pooled effect after correcting for multiple testing due to future living meta-analysis updates (MA1: Hedges’ *g* = −0.225, MA2: Hedges’ *g* = −0.049, MA3: Hedges’ *g* = 0.074). The results were supported by robust Bayesian hierarchical meta-analyses (hBMA), which showed no convincing evidence for an effect for MA1 through MA3 (inclusion BFs 0.692, 0.105, and 0.127). The hBMA for MA4 yielded a Bayes factor of 1.823, indicating anecdotal evidence favoring the alternative hypothesis, thus consistent with the frequentist result and supporting the presence of an effect.

The non-significant results from meta-analyses one to three were further explored with equivalence testing. For MA1 and MA3, the tests yielded non-significant results, and thus we could not establish statistical equivalence for these two meta-analyses. This means that the effect sizes could not be shown to fall within the range of the prespecified equivalence bounds of ± 0.06 and ± 0.23, so they cannot be considered ‘equivalent’ to no effects [97]. For MA2, the equivalence test was non-significant for the bounds of ± 0.06. However, equivalence could be demonstrated when using the less stringent bounds of ± 0.23 (i.e., the effect sizes can be considered ‘equivalent’ to no effects). Reasons for the *non*-significant equivalence test outcomes can be for instance high heterogeneity or low power, which we observed for all analyses. All studies were statistically underpowered to reliably detect a range of effect sizes. In MA1, statistical power was estimated to be 17%, and 6% in MA2 and MA3 (***Figure 4***). MA3 was also the only meta-analysis for which Egger’s regression test was significant and the contour-enhanced funnel plot displayed asymmetry, which is indicative of small study bias and publication bias. Small studies often have low power. Underpowered, small studies with non-significant results tend to be published less frequently [98]. Besides, except for MA4, all meta-analyses were marked by considerable heterogeneity. The observed heterogeneity stemmed predominantly from within-study heterogeneity for MA1 (78%) and MA2 (57%), and from between-study heterogeneity for MA3 (59%). Considering these large heterogeneity estimates, it appears expedient that trial protocols become more consistent. Standardized guidelines with recommendations regarding meal type, meal timing, and oxytocin dosing should be devised, which researchers can follow.

Moderator analysis revealed that intranasal oxytocin dosage had a meaningful impact on the summary effect size estimate for MA2. Populations (with obesity and SCZ) that received 24 IU of oxytocin were associated with a reduction in caloric consumption. In populations (with anorexia, bulimia, and PWS) receiving more than 24 IU relative to 24 IU on the other hand, exogenous oxytocin was linked to less of a decreasing effect on calorie consumption. It is currently still matter of debate what an optimal dose of oxytocin is [108–112], and whether intranasal administration is an adequate administration route [113, 114]. Individuals with PWS, bulimia and anorexia present with widely varying symptoms (i.e., excessive food intake and above-typical weight versus excessive food intake and commonly a typical weight versus reduced intake and below-typical weight). Therefore, it is intriguing that the same higher dose of oxytocin appears to be associated with a similar direction and magnitude of effect. Future studies would benefit from evaluating a range of dosages to ascertain this moderating, possibly context-sensitive effect. Importantly, given the distribution of clinical conditions in the different dosage categories (i.e., 80% of the cohorts that received 24 IU had obesity, >24 IU was received by individuals with anorexia, bulimia, and PWS), it is possible that the moderating effect of the dosage is in fact due to the underlying clinical condition. Furthermore, varying numbers of male and female participants may contribute to observed heterogeneity. Oxytocin is known for acting in a sexually dimorphic manner [115, 116], yet studies are often conducted in only men or only women. Indeed, for MA2, we found a significant moderating effect of the percentage of females in a study cohort on the outcome (*p* = 0.040). Studies with a higher percentage of female participants showed a tendency to have less anorexigenic effects, while studies with only male participants tended to have stronger anorexigenic effects. Of note, for the studies on PWS populations and two populations with obesity, the gender of the analysis group was not clearly reported [38, 61, 62]. The percentage of female participants was 100% in all eating disorder populations present in MA2 (i.e., bulimia nervosa, anorexia nervosa [63–65]), 86% in functional dyspepsia [72], 50% in SCZ [68], and 0% in the remaining population with obesity [34]. It is therefore possible that the population condition was the ‘true’ moderator, rather than the gender.

Generally, the results suggest that oxytocin seems to reduce appetite and related measures (i.e., hunger, satiety, and hedonic drive to eat) in clinical populations with obesity, T2D, SCZ, diabetic gastroparesis, and bulimia nervosa and/or BED (measured in one combined cohort [39]). This result was robust against publication bias and was among the best-powered outcomes. Most of the included conditions are characterized by either atypical, elevated body weight (e.g., obesity, T2D [117], SCZ [118], BED [119]), increased caloric consumption (e.g., bulimia (episodically [120]), obesity [121], T2D [117, 122], BED [119]), or both. It is plausible that oxytocin has an anorexigenic effect on appetite in populations with atypical, excessive body weight and caloric consumption. This is in line with a previous study suggesting that a decrease in food intake through oxytocin is more evident in men with obesity as compared to typical-weight men [34]. In healthy populations, oxytocin might not influence appetite as noticeably, as we also found in the corresponding meta-analysis. However, this result has to be interpreted with caution due to the non-significant equivalence test, which indicates this particular meta-analysis was inconclusive.

Regarding the effect on caloric intake in both healthy and clinical populations, oxytocin showed a marginal effect on reducing caloric intake, which was on the threshold of statistical significance for MA1 after correction for future multiple tests. Non-significant equivalence tests indicated that these results were inconclusive for MA1, and for MA2 within more stringent equivalence bounds. When applying more lenient bounds of ± 0.23, the non-significant summary effect size from MA2 could be considered equivalent. A post-hoc moderator analysis revealed that the type of condition (i.e., whether a population was diagnosed with obesity, anorexia, SCZ, etc.) had a significant moderating effect on the summary effect size (*p* = 0.037). As the only significant moderator level, groups with obesity showed stronger negative effect sizes. This indicates that at least in populations with obesity, oxytocin could be associated with a reduction in caloric intake, too, which would be in line with its curbing effect on appetite in this same population. As briefly mentioned above, the percentage of female participants significantly moderated the summary effect of MA2. A higher percentage of women was linked to a more positive effect. Of note, in this moderator analysis, the anorexia and bulimia groups were all female, and the obesity group was all male. It is therefore conceivable that gender is not the most important moderator that predicts the direction of the effect, but rather that gender is covering a latent, true variable, such as the body weight that is typical for the condition of the participants. A similar mechanism might apply to the significant moderator analysis assessing the covariate ‘oxytocin dose’. Perhaps, the closer to a typical or below-typical body weight, the less oxytocin decreases caloric intake. This explanation appears more likely as it is in line with the results of the corresponding moderator analysis on the condition type. While it is possible that oxytocin acts differently in males and females within the same condition and that different dosages have different effects, this cannot be compared across conditions which have the opposite and widely heterogeneous symptoms and etiologies as hallmark signs (i.e., obesity, anorexia, bulimia, SCZ). Considering that the two isolated effects on caloric intake in bulimia [64] pointed to opposite effects, the role of oxytocin on calorie consumption in this disorder is less clear and would warrant further investigation.

Taken together, exogenous oxytocin seems to be associated with a reduction in appetite and an increase in satiety and fullness in clinical participants, predominantly with conditions typically accompanied by above-typical weight and caloric consumption (e.g., obesity, T2D [117], BED [119]), but not in others (i.e., PWS, anorexia nervosa). Oxytocin may also be linked to a reduction in caloric intake in individuals with obesity. In healthy populations, oxytocin does not seem to have a pronounced anorexigenic effect on appetite and caloric intake. A speculative explanation for this result is that oxytocin could be a *conditional*, context-sensitive [123] anorexigenic that, depending on the underlying disorder, could facilitate a decrease in appetite and caloric intake, thereby promoting the return to a healthy, neutral energy balance and metabolic homeostasis. Oxytocin has been suggested to be involved in regulating metabolic homeostasis before [27]. In populations where there is already a neutral or even negative energy balance or deviation from metabolic homeostasis, the anorexigenic function of oxytocin might be weaker or not present at all. Previously, it has been mostly reported that oxytocin has either no effect on caloric intake and appetite [39, 62, 72] or an anorexigenic effect (e.g., [33, 34, 36, 37].

The results discussed in this systematic review and meta-analysis come with some methodological considerations that should be taken into account. We suggest that oxytocin’s impact on caloric intake and appetite could be context-sensitive and more effective in populations with above-typical body weight and caloric consumption, respectively. We could, however, not test this hypothesis directly in a moderator analysis since most studies did not report their participants’ BMI, body weight or other measures of body composition, and none reported long-term eating habits. It could be that studies that do not report body composition may not have been designed to assess oxytocinergic effects on calorie intake or appetite, which might bias the results. Prospective studies should report these important measures so that they can be incorporated as moderators in a future version of this living meta-analysis.

Furthermore, prospective studies may seek to confirm the effect of exogenous oxytocin on PWS, bulimia nervosa and anorexia nervosa. Regarding PWS, oxytocin did not have a significant effect on caloric intake (see ***Figure 2***). The condition presents itself with symptoms comparable to obesity and BED, which renders this finding intriguing. However, disease diathesis mechanisms and pathways may vary. It is possible, for example, that exogenous oxytocin has a stronger effect on environmentally and behaviorally mediated conditions than genetic ones, such as PWS. On another note, oxytocin receptor expression, oxytocin production, and (downstream) signaling might be altered in PWS populations [124–127], so that receptivity to oxytocin could be affected in individuals with PWS [128]. In bulimia nervosa as an isolated condition (without BED [64]), the effects were mixed. Bulimia is a condition that, in itself, is heterogeneous, with episodes of restriction and binge eating [120]. In anorexia nervosa, the intended treatment effect would be to increase appetite and caloric intake in order to support the return to a healthy body weight. Adaptation to chronic starvation may result in increased appetitive signaling of redundant appetite pathways, which might overcome the anorexigenic effects of oxytocin administration [129]. Currently, there is still a lack of efficacious treatments for anorexia nervosa [130]. At the same time, anorexia is associated with a considerably increased mortality rate [131]. Therefore, novel interventions are required. Aside from that, we observed that most studies included in this meta-analysis were considerably under-powered and highly heterogeneous. Besides the above-mentioned factors (i.e., study design, gender), varying oxytocin formulation can introduce heterogeneity. Studies using a formulation that meets official requirements will likely perform more consistently than ‘home-made’ formulations. Underpowered analyses increase the likelihood of false-negative results [132] and also introduce the chance of a ‘winner’s curse’ if they detect a statistically significant result [133]. Given the insensitive equivalence tests, residual uncertainty remains regarding conclusiveness of the results. Additional well-powered studies of exogenous oxytocin’s effects on caloric intake and appetite are therefore needed.

Another methodological aspect is that the publication bias assessment has not been yet verified for hierarchical, multi-level robust Bayesian model-averaged meta-analysis with the `RoBMA` package [103]. Given, however, that we assessed the presence of publication bias for the frequentist analyses, using a sensitivity analysis [102], Egger’s regression test [102, 104], and contour-enhanced funnel plots [105], the hBMA approach nevertheless seemed valuable, also without a Bayesian publication bias framework. Lastly, this study deviated from the pre-registered protocol in other respects due to resource constraints, unavailability of certain analysis specifications, and the natural structure of the data. All protocol deviations are reported and explained in the supplementary information (supplementary document 1), and directly in respective text sections where appropriate. Post-hoc analyses were clearly declared. The first living meta-analysis update will prioritize performing the remaining two meta-analyses on biomarkers of energy regulation and measures of body composition. This will help bridge the gap between behavioral and physiological metabolic effects of oxytocin. Under the premise that the hBMA will be further developed, the Bayesian analysis will be repeated with publication bias assessment. The analysis scripts and data used for this study are publicly available (see *Data and code availability*), which makes updating and reproducing the meta-analysis accessible and feasible.

In summary, this preregistered, living systematic review and meta-analysis provides evidence indicating that oxytocin may regulate appetite and caloric intake in some clinical populations. This pattern could be explained by homeostatic and anorexigenic properties of oxytocin, which foster energy balance and maintenance in clinical groups with above-maintenance body weight, appetite and/or caloric consumption. This provides further support for a potential therapeutic role of oxytocin in these groups in regulating appetite and other behavioral metabolic markers. Future research should further investigate the mediating role of contextual variables, especially body composition and food intake habits, on the metabolic and energy regulatory effects of oxytocin.

## Data Availability

Data used for analyses are available online at https://osf.io/3zuc8/

## Data and code availability

Supplementary information, data and scripts used for analyses with information on specific parameter settings and additional notes are available at https://osf.io/3zuc8/.

## Acknowledgments

This research was funded by Research Council of Norway (301767). We thank Glenn Karlsen Bjerkenes from the University of Oslo Library for his assistance with determining the databases and search terms for the literature search. All methods were performed in accordance with the relevant ethical guidelines and regulations.

## Author contributions

AIS, DSQ and EAL conceived and planned the study. AIS screened the studies, with contributions from ED and DSQ. Data was extracted by AIS, with contributions from DD. AIS analyzed the data. DSQ supervised the study. DSQ provided funding for the study. AIS interpreted the results, with contributions from ED, HK, DD, KMW, EAL, KTE, DVDM, LTW, OAA and DSQ. AIS wrote the first and revised drafts of the manuscript, with ED, HK, DD, KMW, EAL, KTE, DVDM, LTW, OAA and DSQ contributing to the first and revised drafts of the manuscript.

